# Incontinence and Healthcare Utilization of Medicare Patients

**DOI:** 10.1101/2022.01.25.22269793

**Authors:** Ian Duncan, Andrew Stocking, Karen Fitzner, Tamim Ahmed, Nhan Huynh

**Affiliations:** Department of Statistics and Applied Probability, University of California Santa Barbara; PBE Inc; Santa Barbara Actuaries Inc

**Author notes:** **Address Correspondence to:** Ian Duncan, Dept. of Statistics & Applied Probability, UCSB, South Hall 5518, Santa Barbara, CA 93106. **Source of Funding:** Funding for this paper was provided by PBE Inc. The Office of Research Human Subjects Department at UCSB reviewed this work and determined that it did not meet the criteria for human subjects research as defined in the Common Rule (45 CFR 46). IRB review and oversight are not required because the activity does not involve “human subjects” as defined under 45 CFR 46.102 and the research team is not accessing identifiable private information.

**Keywords:** Incontinence, prevalence, administrative claims, fee-for-service Medicare, incontinence-related events

## Abstract

**Purpose:** Claims data are used to directly measure the prevalence of incontinence and incontinence-related events within a large Medicare population.

**Design:** Retrospective analysis.

**Subjects and Settings:** The study relied on administrative claims data from the CMS Medicare Limited Data Set (5% sample) in 2018. The analysis was limited to fee-for-service (FFS) Medicare beneficiaries with minimum of 3-month enrollment in Parts A & B and at least 65 years old.

**Methods:** We used diagnosis codes to identify members with incontinence in their 2018 claims experience and grouped these members into 3 categories (urinary incontinent only, fecal incontinent only, and dual incontinent (DI)) and four sites-of-service (nursing home, Skilled-nursing Facility (SNF), home health, and self-care). We then determined the incidence of four types of incontinence-related events for each cohort: 1) Urinary Tract Infections (UTIs), 2) Incontinence-Associated Dermatitis (IAD), 3) Slips and falls, and 4) Behavioral disruptions.

**Results:** We found that 11.2 percent of Medicare members had a claims-based diagnosis of incontinence in 2018. This result falls below the estimated share of the over-65 population who are incontinent, as reported in the literature. The prevalence of the four incontinence-related events is significantly higher (between 2 percentage points to 17 percentage points) for members who experience dual incontinence relative to those with only urinary incontinence. On average, those diagnosed with incontinence experienced 5 times more UTIs, two times as many dermatitis events, more than twice as many slips and falls, and 2.8 times more behavior disruptions compared to those without an incontinence diagnosis.

**Conclusions:** Although we find that the prevalence of incontinence is under-reported relative to the literature, our results show that those who are diagnosed as incontinent experience a much higher prevalence of UTIs, IAD, slips and falls, and behavioral deterioration compared to those who are not diagnosed as incontinent. Our results suggest that incontinence may be an important indicator diagnosis of many other conditions and, if not well-managed, may challenge the desire for those who are incontinent to age at home.

## 1. Introduction

Urinary incontinence (UI) and fecal incontinence (FI) are conditions that affect millions of individuals over the age of 65 in the United States and are reported to have a larger effect on quality-of-life measures than diabetes, cancer, or arthritis [1]. The likelihood of incontinence has been found to increase with age, body-mass index, and diabetes, along with any type of prostrate surgeries or childhood diseases for men or natural births for women [2-5]. With the exception of serious injuries, surgeries, or childhood diseases, incontinence often starts light (small, intermittent losses of urine) and increases in severity over time, particularly if BMI or other co-morbidities also increase over time [6].

For men and women over the age of 65, estimates of UI range up to 60% for women and 35% for men [7]. Fecal incontinence is estimated to affect about a quarter to a third of those with UI over the age of 50 [8]. Urinary incontinence, particularly when paired with FI, is a leading cause of admission to nursing homes and skilled care [2, 9, 10]. Although only about 10% of the Medicare population is aging in an institutional setting (long-term care or skilled nursing facility), an estimated 45% to 60% of those Medicare beneficiaries have UI [2, 9-11]. The prevalence of FI in nursing homes is measured less precisely, but three studies suggest that half to three-quarters of nursing home residents who experience UI may also experience FI with a higher incidence of FI among men [2, 5, 7, 11].

The remaining Medicare population is aging at home under selfcare, the care of a home health agency or has been discharged to hospice care. The prevalence of urinary incontinence in the home setting is estimated to be 46% for female Medicare beneficiaries and 28% for male beneficiaries with about 10% of men and women experiencing both UI and FI [2]. For those under hospice care, the rate of UI is over 60% with almost half of those experiencing both UI and FI. Overall, the literature-estimated value of incontinence prevalence for both men and women in home care and nursing home are 39% and 50%, respectively.

The most common management strategy for urinary incontinence is the use of disposable body-worn absorbent products (BWAP), although surgery, medications, and other types of preventative or collection medical devices are also available. For those using a BWAP, the most common mode of failure is leakage, which can result in several complications for the individual including an increase in slips and falls [12], skin breakdown called Incontinence Associated Dermatitis [13-15], serious wounds or infections including UTIs, and a reduction in sleep quality that can contribute to cognitive decline and behavioral conditions [16]. Incontinence has been associated with both an increased need for care assistance and an increased turnover of caregivers [17, 18].

Despite the emotional distress and financial burden to the individuals and to the society, incontinence is often under-reported [15]. The reported incontinence prevalence is widely varied by studies due to difference in study designs, the condition measurements, methodologies, and sample selection. Our main objective is to measure the prevalence of incontinence and incontinence-related events by type and care settings in the 5% Medicare datasets.

## 2. Data and Methods

This analysis relied on administrative claims data from the CMS Medicare Limited Data Set (5% sample) in 2018, and all results for this paper were generated using SAS software version 9.4. We limited our analysis to fee-for-service (FFS) Medicare beneficiaries (i.e., we excluded Medicare Advantage beneficiaries) because of gaps in claims reporting in the Medicare Advantage (MA) beneficiary segment. To reduce duration bias caused by beneficiaries who have minimal exposure to Medicare, we also required at least 3 months of enrollment in each year per member. Our analysis includes only the over-65 Medicare population, which we find to represent approximately 75% of the FFS beneficiary group. These criteria yielded approximately 1.2 million FFS beneficiaries with coverage in 2018.

To identify urinary and fecal incontinence in the longitudinal sample population, we used ICD-10-CM diagnosis codes captured in their claims experience for utilization analysis. Codes for relevant diagnoses, identified in any position on a claims record, are provided in Appendix A. Incontinent members are grouped into 3 categories using the code list: urinary incontinence only (UI), fecal incontinence only (FI), and dual incontinence (DI), which includes members with both UI and FI. Members are then divided into 4 place-of-service categories: nursing home, skilled nursing facility (SNF), home health, and none/other (selfcare) according to the most resource-intensive site in which they received care during 2018.^1^

We then determine the incidence of four types of incontinence-related events for each cohort: 1) UTIs, 2) Incontinence Associated Dermatitis or Perineal Dermatitis (IAD), 3) Slips, falls and related fractures, and 4) claims associated with a behavioral disruption (See Appendix A for their ICD-10-CM diagnosis codes). Incidence for each event is calculated as the share of members within each cohort who experienced at least one event in the year of the UI/FI diagnosis. We also calculate the conditional average number of events or the average number of events for those members in a cohort who experienced at least one event.

## 3. Results

Summary statistics of those diagnosed as incontinent in 2018 by site-of-service and type of incontinence are presented in Table 1. They are compared to a Control consisting of members in the same place-of-service who were not observed as incontinent in 2018.

**Table 1.** Summary Statistics for Incontinent and Not Incontinent Populations, 2018.

| Metrics | Nursing Home |  | SNF |  | Home Health |  | Home / Self Care |  | TOTAL |  |
| --- | --- | --- | --- | --- | --- | --- | --- | --- | --- | --- |
|  | Inco. Cohort | Not Inco. Cohort | Inco. Cohort | Not Inco. Cohort | Inco. Cohort | Not Inco. Cohort | Inco. Cohort | Not Inco. Cohort | Inco. Cohort | Not Inco. Cohort |
| **Urinary Incontinence (UI)** |  |  |  |  |  |  |  |  |  |  |
| Prevalence | 13.5% of 10,635 |  | 17.3% of 52,168 |  | 21.5% of 160,736 |  | 7.9% of 1,019,634 |  | 10.1% of 1,243,173 |  |
| Female | 71.1% | 70.5% | 65.7%*** | 63.7% | 65.6%*** | 61.6% | 63.0%*** | 54.8% | 64.0%*** | 56.0% |
| Average HCC Score | 2.18** | 1.86 | 3.51*** | 2.95 | 3.17*** | 2.70 | 1.36*** | 1.04 | 2.00*** | 1.28 |
| Average Age | 83.0 | 84.0 | 81.0** | 82.0 | 81.0*** | 80.0 | 76.0*** | 74.0 | 78.0*** | 75.0 |
| **Fecal Incontinence (FI)** |  |  |  |  |  |  |  |  |  |  |
| Prevalence | 0.5% of 10,635 |  | 0.8% of 52,168 |  | 0.9% of 160,736 |  | 0.4% of 1,019,634 |  | 0.5% of 1,243,173 |  |
| Female | 62.9% | 70.5% | 64.9% | 63.7% | 63.8%* | 61.6% | 69.9%*** | 54.8% | 68.2%*** | 56.0% |
| Average HCC Score | 2.52 | 1.86 | 4.06** | 2.95 | 3.51*** | 2.70 | 1.44*** | 1.04 | 2.07*** | 1.28 |
| Average Age | 84.0 | 84.0 | 80.0 | 82.0 | 80.0 | 80.0 | 76.0*** | 74.0 | 77.0*** | 75.0 |
| **Urinary & Fecal Incontinence (DI)** |  |  |  |  |  |  |  |  |  |  |
| Prevalence | 2.6% of 10,635 |  | 2.5% of 52,168 |  | 2.2% of 160,736 |  | 0.3% of 1,019,634 |  | 0.7% of 1,243,173 |  |
| Female | 71.8% | 70.5% | 63.4% | 63.7% | 64.0%** | 61.6% | 76.8%*** | 54.8% | 69.14%*** | 56.0% |
| Average HCC Score | 2.34* | 1.86 | 3.90*** | 2.95 | 3.77*** | 2.70 | 1.71*** | 1.04 | 2.95*** | 1.28 |
| Average Age | 83.0 | 84.0 | 82.0 | 82.0 | 81.0** | 80.0 | 77.0*** | 74.0 | 80.0*** | 75.0 |
| **Total Incontinence** |  |  |  |  |  |  |  |  |  |  |
| Prevalence | 16.6% of 10,635 |  | 20.6% of 52,168 |  | 24.5% of 160,736 |  | 8.6% of 1,019,634 |  | 11.2% of 1,243,173 |  |
| Share of UI with FI | 16.1% of 1,710 |  | 12.5% of 10,329 |  | 9.1% of 37,961 |  | 3.7% of 83,502 |  | 6.1% of 133,502 |  |
| Female | 71.0% | 70.5% | 65.4% | 63.7% | 65.4% | 61.6% | 63.8% | 54.8% | 64.5% | 56.0% |
| Average HCC Score | 2.22 | 1.86 | 3.58 | 2.95 | 3.25 | 2.70 | 1.38 | 1.04 | 2.08 | 1.28 |
| Average Age | 83.0 | 84.0 | 81.1 | 82.0 | 81.1 | 80.0 | 81.0 | 74.0 | 78.1 | 75.0 |

**Note:** Asterisks represent significant difference between the incontinent and not incontinent groups within each place-of-service setting:
‘\*\*\*’ indicates p-value<.0001; ‘\*\*’ indicates p-value<.05; ‘\*’ indicates p-value<.10

### 3.1 Prevalence of Incontinence by Type and Site of Service

Our first research objective was to compare the prevalence of UI and/or FI among the sample beneficiaries compared with the values reported in the literature. In 2018, we find that 11.2 percent of the members were diagnosed with incontinence (urinary or fecal) (Table 1).^2^

As predicted in the literature, we find that the prevalence of incontinence is higher in places-of-service that offer more intensive care: 20.6 percent for SNFs, 16.6 percent for nursing homes, 24.5 percent for those receiving formal healthcare at home compared to 8.6 percent under selfcare. This trend is even more pronounced for those diagnosed with dual incontinence: 62 percent of those with dual incontinence are under some type of formal care, either institutional or home healthcare compared to 36 percent of those with only a urinary incontinence diagnosis.

The results in Table 1, however, identify far fewer incontinent members than reported in the literature. We do not take this as evidence that the literature is incorrect, but instead that CMS claims data on incontinence is incomplete, as found in other datasets [15]. A well-known limitation of claims data, including CMS claims data, is the lack of clinical evaluation. Other studies have gone through pain-staking effort to evaluate large amounts of clinical data to ascertain the prevalence of UI [19-21]. The fecal incontinence results in Table 1 suggest that CMS data is missing many FI individuals. The literature predicts that between half and three-quarters of those in skilled nursing facilities with UI also experience FI: Nelson [19] reports that the prevalence of FI in nursing homes can be as high as 50%; Leung and Schnelle [7] estimate prevalence as high as 65%. Table 1 reports that fecal incontinence comprises only 16 percent of those with UI in nursing homes, 12 percent in SNFs, and 9 percent in home health. Identifying the missing urinary and fecal incontinent members is a topic of other research.

### 3.2 Prevalence of Incontinence-Related Events by Type and Site of Service

We next examined the incidence of incontinence-related events for those observed with incontinence in 2018, for each places-of-service, comparing the incontinent cohort with the cohort not observed as being incontinent (Table 2). In nearly all incontinent group pairs, we observe statistically-significantly higher rates of UTIs, IAD, slips and falls, and behavioral disruptions. These results are consistent with the literature in showing a connection between incontinence and the selected incontinence-related events. Considering the 2018 population, those with urinary incontinence are three times more likely to have at least one UTI and, conditional on having a UTI, will have 1.6 more UTIs than someone who has not been identified as incontinent. The difference increases for those with dual incontinence, who are almost four times more likely to have at least one UTI and, conditional on having a UTI, will have more than twice as many UTIs as someone who has not been diagnosed as incontinent.

**Table 2.** Summary of 2018 Urinary & Fecal Incontinent Groups by Place-of-Service.

| Metrics |  | Nursing Home |  | SNF |  | Home Health |  | Home / Self Care |  | TOTAL |  |
| --- | --- | --- | --- | --- | --- | --- | --- | --- | --- | --- | --- |
|  |  | Inco. Cohort | Not Inco. Cohort | Inco. Cohort | Not Inco. Cohort | Inco. Cohort | Not Inco. Cohort | Inco. Cohort | Not Inco. Cohort | Inco. Cohort | Not Inco. Cohort |
| **Urinary Incontinence (UI)** |  |  |  |  |  |  |  |  |  |  |  |
| UTIs | Incidence | 42.6%*** | 26.0% | 60.5%*** | 38.4% | 54.5%*** | 30.3% | 30.2%*** | 9.4% | 39.2%*** | 12.9% |
|  | Cond. Average | 3.34*** | 2.66 | 6.16*** | 4.30 | 5.67*** | 3.71 | 2.88*** | 1.96 | 4.31*** | 2.69 |
| Dermatitis | Incidence | 7.9%*** | 4.4% | 10.7%*** | 6.4% | 9.2%*** | 6.2% | 6.7%*** | 4.1% | 7.7%*** | 4.5% |
|  | Cond. Average | 2.02*** | 1.75 | 2.21*** | 2.04 | 1.80*** | 1.67 | 1.47*** | 1.41 | 1.66*** | 1.49 |
| Slips & Falls | Incidence | 47.1%*** | 39.6% | 66.4%*** | 56.2% | 71.7%*** | 60.8% | 31.6%*** | 22.2% | 45.3%*** | 27.8% |
|  | Cond. Average | 4.54*** | 3.81 | 7.10*** | 6.29 | 7.25*** | 6.17 | 3.49*** | 3.15 | 5.52*** | 4.12 |
| Behavior Disruptions | Incidence | 19.9% | 19.9% | 21.3% | 21.5% | 10.2%*** | 6.8% | 1.3%*** | 0.6% | 5.4%*** | 2.2% |
|  | Cond. Average | 6.02** | 5.29 | 6.39 | 6.29 | 4.75*** | 4.46 | 3.16*** | 2.98 | 5.03*** | 4.88 |
| **Fecal Incontinence (FI)** |  |  |  |  |  |  |  |  |  |  |  |
| UTIs | Incidence | 32.1% | 26.0% | 52.3%*** | 38.4% | 45.0%*** | 30.3% | 18.3%*** | 9.4% | 26.8%*** | 12.9% |
|  | Cond. Average | 3.41 | 2.66 | 4.81** | 4.30 | 5.05*** | 3.71 | 2.35*** | 1.96 | 3.73*** | 2.69 |
| Dermatitis | Incidence | 0.0% | 4.4% | 9.30%*** | 6.4% | 10.04%*** | 6.2% | 6.83%*** | 4.1% | 7.68%*** | 4.5% |
|  | Cond. Average | 0/00 | 1.75 | 2.87*** | 2.04 | 1.97*** | 1.67 | 1.58** | 1.41 | 1.80*** | 1.49 |
| Slips & Falls | Incidence | 45.3% | 39.6% | 66.8%*** | 56.2% | 73.7%*** | 60.8% | 33.2%*** | 22.2% | 44.9%*** | 27.8% |
|  | Cond. Average | 7.0** | 3.81 | 6.75 | 6.29 | 6.77** | 6.17 | 3.33** | 3.15 | 5.01*** | 4.12 |
| Behavior Disruptions | Incidence | 11.3% | 19.9% | 19.1% | 21.5% | 9.7%*** | 6.8% | 1.2%*** | 0.6% | 4.5%*** | 2.2% |
|  | Cond. Average | 3.67*** | 5.29 | 5.55** | 6.29 | 4.52 | 4.46 | 3.14*** | 2.98 | 4.53*** | 4.88 |

| **Urinary & Fecal Incontinence (DI)** |  |  |  |  |  |  |  |  |  |  |  |
| --- | --- | --- | --- | --- | --- | --- | --- | --- | --- | --- | --- |
| UTIs | Incidence | 35.5%*** | 26.0% | 54.5%*** | 38.4% | 58.7%*** | 30.3% | 34.0%*** | 9.4% | 47.9%*** | 12.9% |
|  | Cond. Average | 3.68** | 2.66 | 6.34*** | 4.30 | 6.51*** | 3.71 | 3.52** | 1.96 | 5.61*** | 2.69 |
| Dermatitis | Incidence | 6.2% | 4.4% | 11.7%*** | 6.4% | 11.5%*** | 6.2% | 8.2%*** | 4.1% | 9.9%*** | 4.5% |
|  | Cond. Average | 2.59*** | 1.75 | 3.48*** | 2.04 | 2.05*** | 1.67 | 1.66*** | 1.41 | 2.20*** | 1.49 |
| Slips & Falls | Incidence | 57.3%*** | 39.6% | 68.7%*** | 56.2% | 77.0%*** | 60.8% | 43.3%*** | 22.2% | 62.3%*** | 27.8% |
|  | Cond. Average | 5.68*** | 3.81 | 7.40*** | 6.29 | 8.18*** | 6.17 | 4.16*** | 3.15 | 6.91*** | 4.12 |
| Behavior Disruptions | Incidence | 27.5%*** | 19.9% | 27.9%*** | 21.5% | 19.0%*** | 6.8% | 4.9%*** | 0.6% | 15.4%*** | 2.2% |
|  | Cond. Average | 5.17 | 5.29 | 5.93** | 6.29 | 5.15*** | 4.46 | 4.01*** | 2.98 | 5.24*** | 4.88 |

**Note:** '\*\*\*' indicates p-value<0.0001; '\*\*' indicates p-value<0.05; '\*' indicates p-value<0.1

The same pattern emerges for dermatitis, slips and falls, and behavioral disruptions. Those with incontinence, on average, are 72% more likely to have dermatitis, 63% more likely to experience a slip and fall, and more than twice as likely to experience a behavioral disruption compared to those without an observed incontinence diagnosis. And once a member experiences one of these conditions, they, on average, have 12% more dermatitis episodes, 34% more slips and falls, and the same number of behavior disruptions as those without observed incontinence.

Consistent with the literature, we find that members with a higher incidence of UTIs, dermatitis, slips and falls, or behavioral disruptions are cared for by places-of-service with more intensive care [21,22]. For example, in the most intensive place-of-service, SNFs, 60.5 percent of incontinent members in 2018 had at least one UTI with a conditional average of more than six UTIs in 2018. This is compared to 30.2 percent of incontinent members who are aging at home under self-care with a conditional average of 2.9 UTIs in 2018. Similarly, in 2018, 66.4 percent of incontinent members in a SNF had at least one slip or fall compared to 31.6 percent of incontinent members aging at home.

## 4. Discussion

This is the first paper to use Medicare claims data to directly measure the prevalence of incontinence. We found that 11.2 percent of the members were diagnosed with incontinence in 2018. This percentage falls below the share estimated in the literature of the over-65 population who are incontinent. We hypothesize that the incontinent population identified through claims in the CMS data is likely to miss many of those with light incontinence because light incontinence is less likely to be diagnosed by a clinical professional and less likely to be identified as a comorbidity when compared to more severe incontinence [22].

We have a rich set of Medicare claims that allows us to quantify the incidence of incontinence-related events and comorbidities within the incontinent population, as compared to the non-incontinent population. The prevalence of UTIs, dermatitis, slips and falls, and behavioral deterioration increase by a statistically significant amount for those with incontinence compared to those without. On average, those diagnosed with incontinence will, in the year of diagnosis, experience 5 times more UTIs, two times as many dermatitis events, more than twice as many slips and falls, and 2.8 times more behavior disruptions compared to those without incontinence.

We observe that the incidence of the four incontinence-related events is significantly higher for those who experience dual incontinence relative to those with only urinary incontinence. For example, the incidence of a UTI in 2018 increases from 39 percent to 48 percent for those diagnosed with dual incontinence compared to those with just a UI diagnosis. Similarly, the incidence of dermatitis in 2018 increases from 8 percent to 10 percent; incidence of slips and falls increases from 45 percent to 62 percent; incidence of behavior disruption in a nursing home increases from 5 percent to 15 percent. These results are consistent with the significant increase in risk posed by the combination of feces and urine to skin health [13].

### 4.1 Limitations and Considerations

Our approach in using Medicare claims data to assess the prevalence of incontinence is subject to several limitations. First, our database does not contain any drug records, although this may not be a significant limitation as incontinence is not commonly treated with medication. Second, we identify patients through administrative claims and thus we do not have access to clinical information to determine members’ diagnosis and severity. Third, our recorded incontinence prevalence is less than that which has been estimated in the literature, suggesting that incontinence is either not sufficiently severe to result in a formal diagnosis or physicians are not incented to code the incontinence diagnosis. Under-coding is well-known in Medicare fee-for-service compared with Medicare Advantage (for example [23, 24]). Finally, we note that our analysis does not allow us to claim that incontinence is the cause of the comorbidities analyzed, but only that there is a correlation between incontinence and the comorbidities.

## 5. Conclusions

The incontinent population is associated with disproportionately high incidence of UTIs, dermatitis, slips and falls, and behavioral disruptions, relative to those without an incontinence diagnosis. The high frequency of these incontinence-related events can have serious negative health and wellbeing effects for those with incontinence and may cause a change in the place-of-service to one of increasing intensity of care. In addition, each event increases the level of effort required of the caregiver.

Even before the COVID pandemic, trends in the United States were to age at home because of lower costs and increased comfort for the aging population. Aging at home also reduces the burden of staffing for institutional settings like nursing homes and skilled nursing facilities, which already have difficulty attracting and retaining staff [25]. However, aging at home shifts the burden of care to friends, family, and private care staff who are often less prepared for healthcare complications. As described in the literature and reinforced by the results of this paper, incontinence and particularly dual incontinence is highly correlated with conditions that require more intensive care and may trigger a transition to nursing or skilled nursing care. Consequently, successfully managing incontinence would seem to be a critical precursor to supporting the desire to age at home.

## Data Availability

All data produced in the present study are available upon reasonable request to the authors

## Acknowledgment

The authors thank Elaine Zhao ASA MAAA and Maggie Richard FSA MAAA for research assistance.

## Appendix A.

### ICD-10 Diagnosis Code List for Incontinence and Related Conditions

Appendix A. ICD-10 Diagnosis Code List for Incontinence and Related Conditions
| Condition | ICD-9 Diagnosis Code | ICD-10 Diagnosis Code |
| --- | --- | --- |
| Urinary Incontinence | 788.30 | R32 Unspecified urinary incontinence |
|  | 788.63 | R39.15 Urge Incontinence |
|  | 788.91 | R39.81 Functional urinary incontinence |
|  | 596.54, 596.59 | N31.9 Neurogenic Bladder |
|  | 596.51 | N32.81 Overactive bladder |
|  | 596.6, 596.7, 596.89 | N32.89 Bladder spasm |
|  | 625.6, 788.32 | N39.3 Stress incontinence (female) (male) |
|  | 788.31 | N39.41 Urge incontinence |
|  | 788.34 | N39.42 Incontinence without sensory awareness |
|  | 788.33 | N39.46 Stress and Urge incontinence |
|  | 788.36 | N39.44 Nocturnal urinary Incontinence |
|  | N/A | N39.492 postural (urinary) incontinence |
|  | 788.39 | N39.498 reflex urinary incontinence |
| Fecal Incontinence | * | R15 Fecal incontinence |
|  | * | R15.0 Incomplete defecation |
|  | * | R15.1 Fecal smearing |
|  | * | R15.2 Fecal urgency |
|  | * | R15.9 Full incontinence of feces |
|  | * | R98.1 Fecal incontinence, nonorganic origin |
|  | * | K59.2 Neurogenic Bowel |
| Urinary Tract Infections | 599.0 | N39.0 Urinary tract infection, site not specified |
|  | 595 | N30 Cystitis |
|  | 597.0, 099.4, 597.8 | N34 Urethritis and urethral syndrome |
| Incontinence Associated Dermatitis or Perineal Dermatitis | 691.0 | L22 Diaper dermatitis |
|  | N/A | L30.9 Dermatitis, unspecified |
|  | N/A | L24.9 Irritant contact dermatitis, unspecified cause |
|  | 112.3 | B37.2 Candidiasis of skin and nail |
|  | 112.1 | B37.3 Candidiasis of vulva and vagina |
| Slips, Falls, and Fractures | N/A | R29.6 Repeated Falls |
|  | V15.88 | Z91.81 history of falling |
|  | E880-E884, E885.9, E886.0, E887-E888, E917.7, E917.8, E929.3 | W00 – W19 Slipping, tripping, stumbling and falls |
|  | 800-826, 830-908, 910-918, 921-929, 950-957, 959, V54.1 | S00 – S99 (Injuries of external causes) |
| Behavioral Disruptions | 294.11 | F02.81 Alzheimer's dementia late onset with behavioral disturbance |
|  | 294.21 | F03.91 Unspecified dementia with behavioral disturbance |
|  | 331.19 | G31.09 Dementia with Parkinsonism plus F02.81 dementia with behavioral disturbance |
|  | 306.8 | F59 Unspecified behavioral syndromes associated with physiological disturbances and physical factor |
|  | 312.9 | F91.9 Conduct disorder |

Note: \* Used ICD9 codes for the analysis of latent incontinence and did not evaluate fecal incontinence

## Footnotes

1 CMS considers a Skilled-nursing Facility to be a facility that primarily provides inpatient skilled nursing care and related services to patients who require medical, nursing, or rehabilitative services, but does not provide the level of care or treatment available in a hospital. A nursing facility can do the same in addition to offer, on a regular basis, health-related care services above the level of custodial care to other than individuals with intellectual disabilities. See: https://www.cms.gov/medicare/coding/place-of-service-codes.

2 We analyzed data from the 2014-2017 period and found similar results as presented here for 2018.

